# SimCOVID: Open-Source Simulation Programs for the COVID-19 Outbreak

**DOI:** 10.1101/2020.04.13.20063354

**Authors:** Ismael Abdulrahman

## Abstract

This paper presents open-source computer simulation programs developed for simulating, tracking, and estimating the COVID-19 outbreak. The programs consist of two separate parts: one set of programs built in Simulink with a block diagram display, and another one coded in MATLAB as scripts. The mathematical model used in this package is the SIR, SEIR, and SEIRD models represented by a set of differential-algebraic equations. It can be easily modified to develop new models for the problem. A generalized method is adopted to simulate worldwide outbreaks in an efficient, fast, and simple way. To get a good tracking of the virus spread, a sum of sigmoid functions was proposed to capture any dynamic changes in the data. The parameters used for the input (infection and recovery rate functions) are computed using the parameter estimation tool in MATLAB. Several statistic methods were applied for the rate function including linear, mean, root-mean-square, and standard deviation. In addition, an adaptive neuro-fuzzy inference system is employed and proposed to train the model and predict its output. The programs can be used as a teaching tool and for research studies.

## 1.1 Introduction

Coronavirus disease 2019 (well-known as COVID-19 or 2019-nCoV) is a disease caused by a novel virus called SARS-CoV-2 [1]. The first reported case of this disease was on Dec. 31, 2019, in Wuhan, China. The outbreak has been declared as a public health emergency of international concern by the World Health Organization (WHO) on Jan. 30, 2020 [2], and as a pandemic on March 11 [3]. The virus spread rapidly around the world and several large-size clusters of the spread have been observed worldwide [4]. According to Johns Hopkins University, the total number of confirmed cases surpassed one million cases on April 2, 2020 [5].

An essential part of minimizing the spread of the virus is to monitor, track, and estimate the outbreak. This is extremely useful for decision making against the public health crises [6]. One way to predict the dynamic spread of the epidemic is by using computer simulation following the mathematical model of an epidemic. In the literature, several analytical approaches have been proposed to model the pandemic including Susceptible-Infected-Removed (SIR) model [6-7], Susceptible-Exposed-Infected-Removed (SEIR) model [1], Susceptible-Infected-Recovered-Dead (SIRD) model [8,9], fractional-derivative SEIR [10], and SEIRD [11]. While some recent studies are addressing this pandemic by developing simulation codes [6-11], there is an increasing need to develop open-source computer programs to perform a time-domain simulation of the dynamic spread of the virus. References [12-13] present MATLAB code scripts to achieve this objective. Reference [14] presents a Python-based program called CHIME (COVID-19 Hospital Impact Model for Epidemics) for hospital uses.

It is advantageous to have an educational program that displays the mathematical model of the virus spread in a visualized block diagram in addition to coded scripts. One of the widely-used platforms for studying the dynamic behavior of a system is MATLAB\Simulink. It has been used by many academic researchers in different fields for simulating dynamic systems using time-domain simulations. A dynamic system, such as the COVID-19 spread, can be mathematically modeled by a set of differential equations (DEs) or a set of differential-algebraic equations (DAEs) depending on the employed model. The main challenge with such a simulation is to estimate the parameters in the model–for instance, the infection and recovery rates. For the SIR model, these two parameters are the inputs to the model, whereas the outputs of the model are the state variables of the system. Since we have initial data to compare (reported confirmed cases of infection), we can estimate the parameters of the infection and recovery rates so that the two outputs (reported and simulated cases) are equal and their difference is near zero. However, parameter estimation could fail in many situations [12–13] due to restricted limits applied to the parameter, unknown initial values of the parameters, and sudden changes in the reported data.

One observation from the proposed models in the literature is that the exponential function is mathematically the major part of the estimation. This is because the problem is nonlinear and the exponential function can represent any increasing or decreasing rates depending on the sign and magnitude of the exponent. A challenge emerges when there are multiple dynamic changes in the reported-cases data or another wave of the pandemic that the simulation struggles to accurately track these data and estimate the future outbreak. This requires variable inputs with multiple step functions to vary its parameters at each time point the data changes its rate dramatically. By applying this strategy, the program solves the problem even when the infection data changes its rate at various levels.

With several open-source programs, this paper introduces a generalized method to track and estimate the virus spread worldwide in an easy, efficient, and fast way. The proposed method is implemented using the SIR and SEIR models and coded under the MATLAB\Simulink platform. The time points where the data changes dynamically are extracted from the data automatically based on some statistics measures. The number of exponential branches for the rate functions are left for the user to choose since each data has its own needs of parameters. The data is taken from [15] and is updated daily for all countries worldwide. The user enters the country name and runs the model with optional editable settings. With the application of the adaptive neuro-fuzzy inference system (ANFIS), the outbreak of China and Italy are implemented in Simulink using both standard mathematical model and ANFIS system.

## 1.2 Introduction to SIR, SEIR, and SIRD models

The SIR model is a basic representation used widely to describe a disease spread, and it is the fundamental model for the other models such as SEIR and SIRD. SIR model consists of three-compartment levels: Susceptible (S), Infectious (I), and Removed (R). Any individual belongs to one of these groups. A brief description of these compartments is given below. Susceptible individuals are those people who have no immunity to the disease but they are not infectious. Since there is no vaccine yet developed for this disease, we can say that the entire community is exposed to get infected by this disease and hence, the “Susceptible” compartment can be represented by the entire population. An individual in the “Susceptible” level can move into the next level of the model (Infectious) through contact with an infectious person. By this single transmission, the number of susceptible\infectious people reduces\increases by one, respectively. The next compartment is for the infectious people who have the disease and can spread it to susceptible people. Infectious people can move to the “Removed” compartment by recovering from the disease. The removed compartment includes those who are no longer infectious and the ones who have dies from the disease (closed cases). The summation of these three compartments in the SIR model remains constant and equals the initial number of population. A basic SIR model is shown in Fig. 1, where *β* denotes the infection rate or the transmission rate, and *γ* denotes the recovery or removed rate. Generally speaking, these parameters *(β, γ*) are not constant; they are functions of the size of infectious and recovery compartments. These are the parameters that we want to optimize and estimate so that the reported and simulated cases are approximately equals. To solve this set of differential equations, we need initial values for the three-state variables *S, I*, and *R* namely *S*_(0)_, *I*_(0)_, and *R*_(0)_. The initial value *S*_(0)_ is the community population impacted by the outbreak [6], whereas, *I*_(0)_ is the number of confirmed cases that can be any value but not zero. We can set *R*_(0)_ to zero, if the start times of the spread and simulation are equals. The transmission rate reduces monotonically with time [6].

**Fig 1b.**
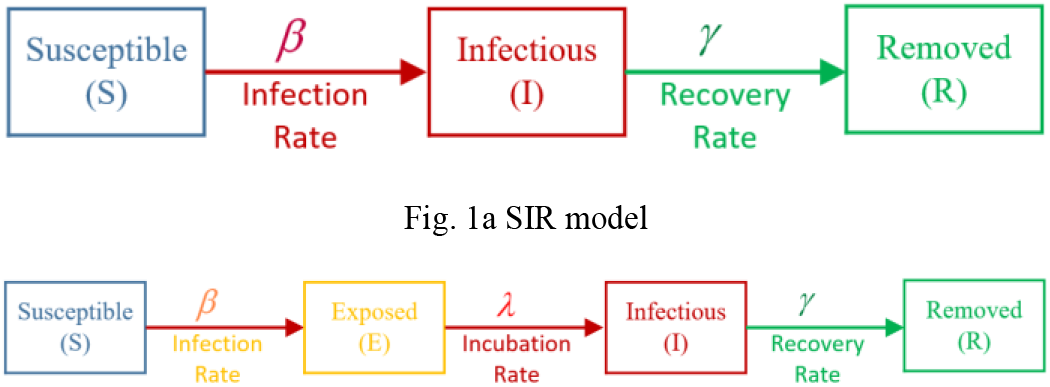
SEIR model

Mathematically, a standard SIR model can be represented using the following differential equations:

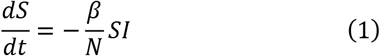

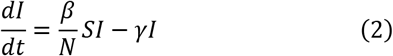

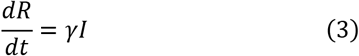

Where *N* denotes the total population size, *N = S + I + R* and the natural birth and death rates are ignored.

For the SEIR model, an additional compartment is added between the Susceptible and Infectious compartment called “Exposed”. This compartment is dedicated to those people who are infectious but they do not infect others for a period of time namely incubation or latent period. Note that the summation of the four state-variables at any time stays constant and equals the initial population, and *λ* in the equations is the reciprocal of the incubation period (which can vary between 2 to 14 days). For the SIRD model, an additional compartment is added at the end of the SIR model to distinguish between recovered and death cases in the “Removed” compartment. It is worth mentioning that the 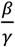 ratio in the SIR model gives us an important metric γ called “basic reproductive number”, or *R*_0_ This ratio is a measure of how contagious a disease is, in other words, how many people are infected by a single infected person. If *R*_0_ > 1, there is a spread of disease which is a strong sign of a pandemic. If *R*_0_ < 1, there is a decline in the spread [16].

## 2. The Proposed Methodology

Since the programs developed in this paper are for simulating any outbreak around the world with potential multiple dynamic changes in the reported cases, we need to look for a generalized method that is applicable for all possible scenarios. The reported data used for comparison is either the daily confirmed\measured infection cases (*I*_m_) or its cumulative infection cases (*C*_m_). These plots are nonlinear curves and can be represented by an exponential formula such as the sigmoid function expressed in (4):

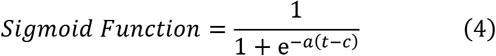

where “*a*” refers to power gain (exponent) which could be positive or negative, and “*c*” refers to a time constant. The above sigmoid function is always positive regardless of the signs of its parameters which satisfies the representation of physical components such as rate variables in this problem. At each time “*c*” when there is a noticeable change in the reported data, the sigmoid function changes its magnitude to find new values for the parameters. In other words, the proposed rate function consists of multiple branches of sigmoid functions; each has different gain and time constant. The final rate function is the aggregation of these branch functions as in expressed in (5).

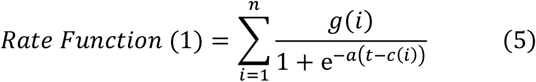

where “*g*” is the gain of branch–*i*.

For simplicity, it is assumed the parameter “*a*” is a constant value to be estimated. The time parameters “*c*” is represented by a vector which its elements can be manually pre-defined or estimated. The length of this vector is the number of sigmoid branches–that is, the number of iterations in (5). The aggregated sigmoid function can be further generalized by subtracting the previous sigmoid function from the one in the consideration. The following rate function is for the latter concept with a generalized infection\recovery rate function used for tracking the problem. The higher the number of branches (*n*), the smoother and better matching between the reported data and the simulated plot.

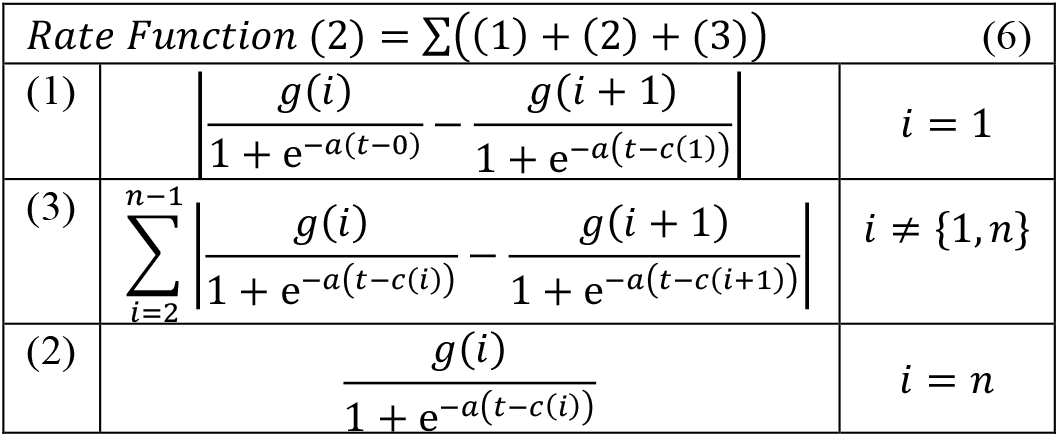

For the estimation problem, we need to multiply the above function with an exponential component having a negative exponent in order to downturn the curve if the pandemic has not yet passed the peak. The parameters of the new component are also estimated by the solver. If the exponent is zero, the function would be as the above equation. However, any nonzero exponent leads to different case studies such as standard, optimistic, and pessimist estimations. The following equation can be used for estimating the parameters with any optional scenario.

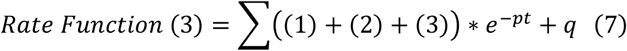

where *p* and *q* are the parameters that need to be estimated.

It should be noted that since the recovery function does not change its rate rapidly–unlike the infection function– due to lack of vaccine at the current time, we can represent this function with only one sigmoid function or even simpler as a constant parameter to be estimated by the program. It is worth mentioning that if *a* = 0 in the aggregated rate function, the sigmoid functions become an aggregated step function. The lower the power gains of the sigmoid function, the steeper the curve is, and vice versa. Figure 2 illustrates the proposed concept assuming different values of *a*, where *S* in the figure refers to a step function.

**Fig 2.**
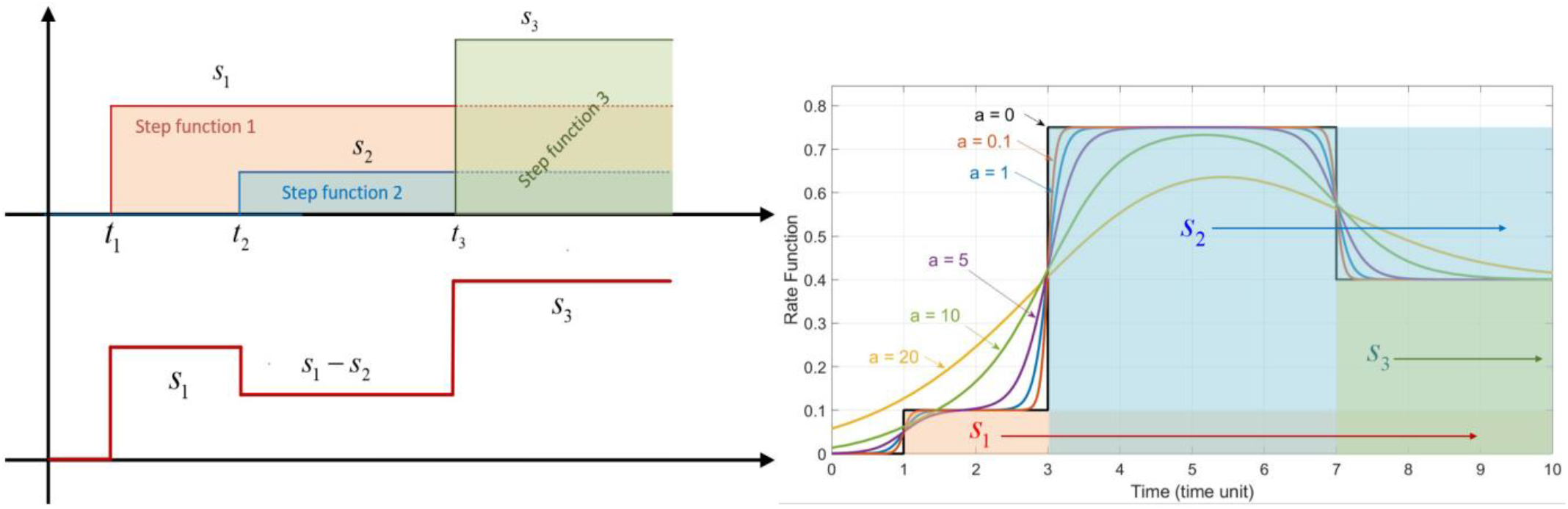
Illustration of the proposed rate function with an aggregated step function (left) and sigmoid function (right).

For the initial values of the parameters, we need a methodology that satisfies all potential outbreaks. One efficient way is to use the confirmed infection ratio 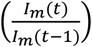 and normalizing the ratio with a range between zero and one. This quantity is employed for the gain parameters of sigmoid branches–that is, *g* in (5). For the time parameters–*c* in (5)– we can use one of these methods: (1) a set of predefined time locations (2) a statistical approach to automatically find the best values for these parameters to optimally fit the curve. Due to its generality and simplicity, the second method is considered as the standard method for the programs. In MATLAB, we have a function called “*MaxNumChanges*” intended to determine some useful statistical information with the following measures: (1) “*linear*” (2) root-mean-square “*r ms*” (3) “*mean*” (4) standard deviation “*std*”. The first technique is considered as the basic tool for the initial time calculation but it can be easily switched to the other techniques by changing its name. Figure 3 illustrates this concept assuming the maximum number of changes 2, 3, 5, 10. Clearly, increasing this quantity provides better output matching and parameter estimation. For many cases, it is found that 3–5 change points (which gives the number of sigmoid branches as well) are sufficient for this problem. However, the choice of the number of change points is left for the user to tune the estimation for each outbreak.

**Fig 3.**
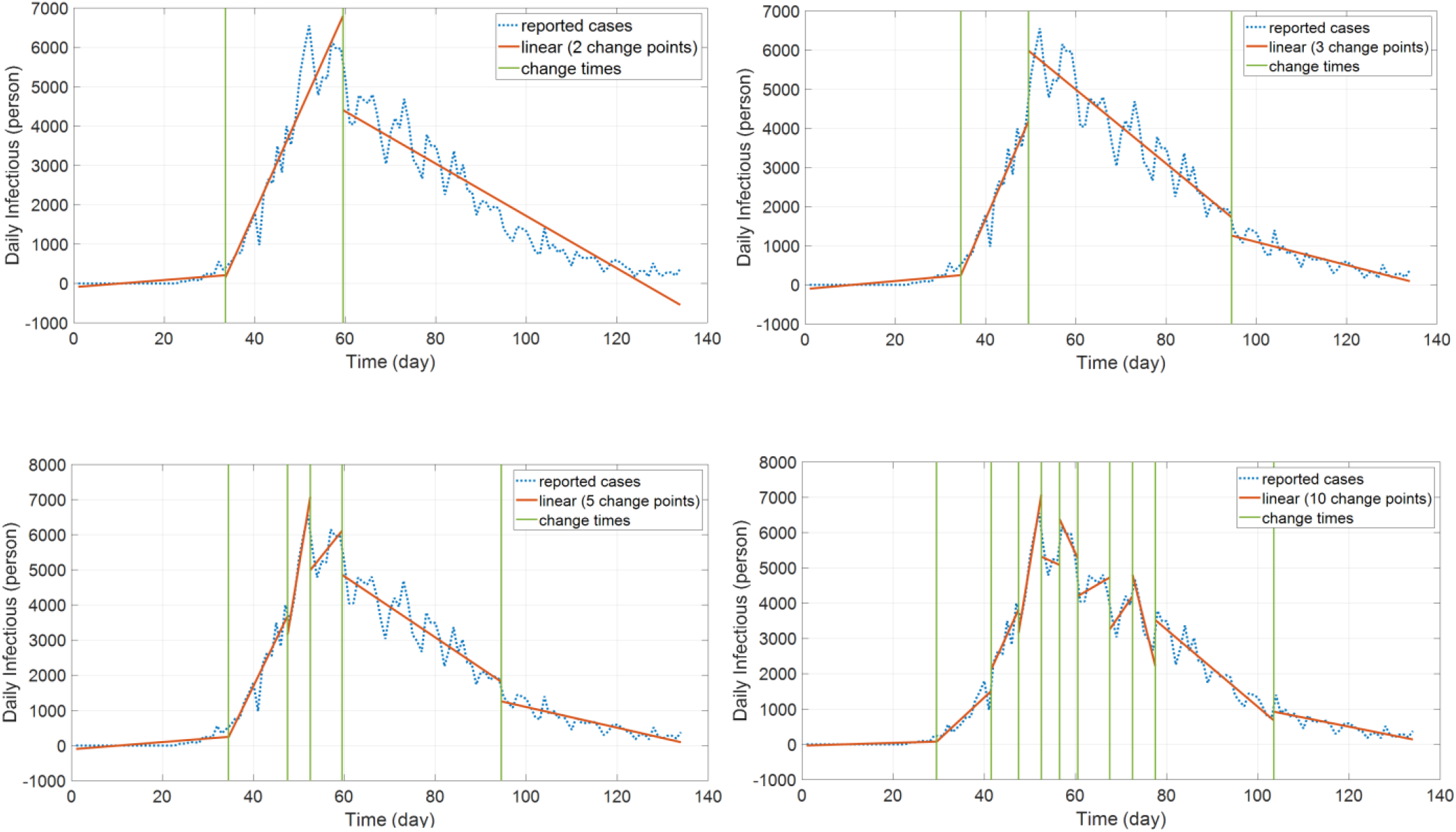
Changing maximum number of change points with values 2, 3, 5, 10, respectively.

**Fig 4.**
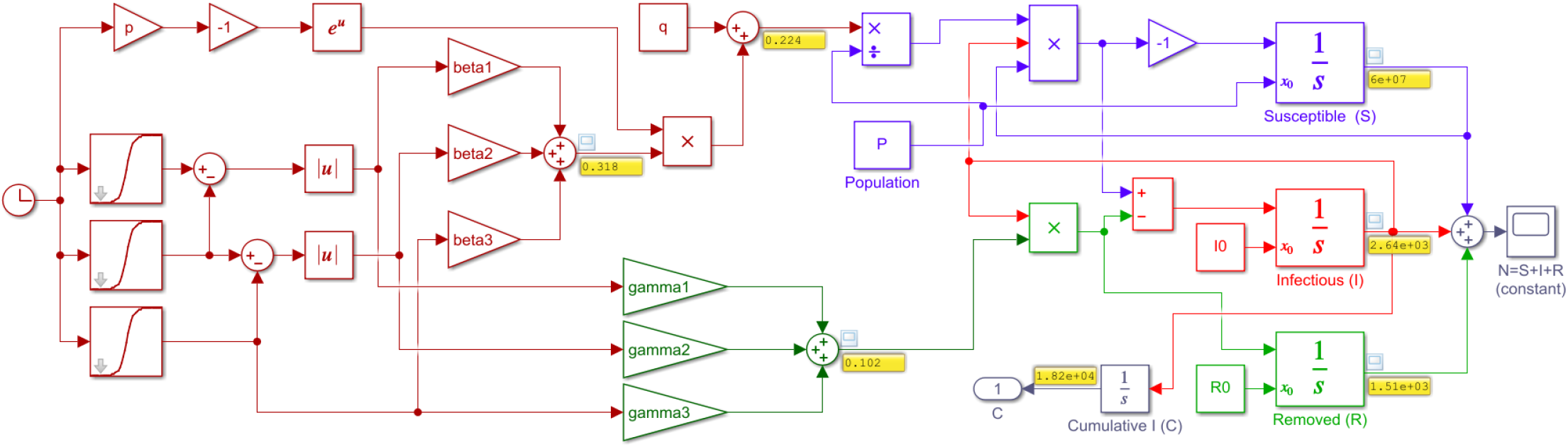
SIR model in Simulink for simulating the virus spread with three sigmoid branches– blue is for “Susceptible” with its input (β), red is for “Infectious”, and green is for “Recovered” with its input (γ)

## 3. Simulation Example

To see how the problem is modeled in Simulink using the differential equations (1)-(3), a simulation example is shown in Fig. 3 with the proposed concept of rate function assuming three sigmoid branches. A similar program is coded in MATLAB. In the diagram, different colors are used to distinguish the equations (blue for “Susceptible”, red for “Infectious”, and green for “Removed” compartment). Note that the control measure parameters (*β, γ*) are functions of time to be found and estimated by the program. All the numerical data for the test cases were taken from [16]. The test outbreaks used in this section are the cases of China, Italy, and Iran–which has several fast-dynamic changes in the data– assuming multiple sigmoid branches in the models. Figure 5 displays part of the results for the outbreak of China using SimCOVID. The plot on the left is for the beta-gamma function which varies over time and provides some useful information about the reproduction number and the overall pandemic. The plot on the right is a two-y-axis plot showing the infection and its cumulative cases. If the number of sigmoid input changes, these plots change as well and exhibit more detail that is hidden in the simple model. The same program is used to plot the graphs in Fig. 6a which is related to the Italy outbreak and the estimation is carried out on April 7^th^, 2020. The estimation ends by the end of July 2020 which shows good matching between the measured cases (released later after this estimation) and the simulation results given in Fig. 6a–b. The gray area in the plots is for confidence interval on the same date of estimation with ±10%. Figure 7 displays part of the results obtained for Iran outbreak and the estimation occurs on June 21^th^, 2020 using the MATLAB codes (added later to the package). This outbreak is chosen due to its multiple dynamic changes in the reported cases to show the capability of the programs to simulate such a problem. As can be observed, the programs can be used for simulating the epidemic assuming various scenarios and confidence intervals at different times. Applying new measures is easy to see the impact of these control efforts on future estimation.

**Fig 5.**
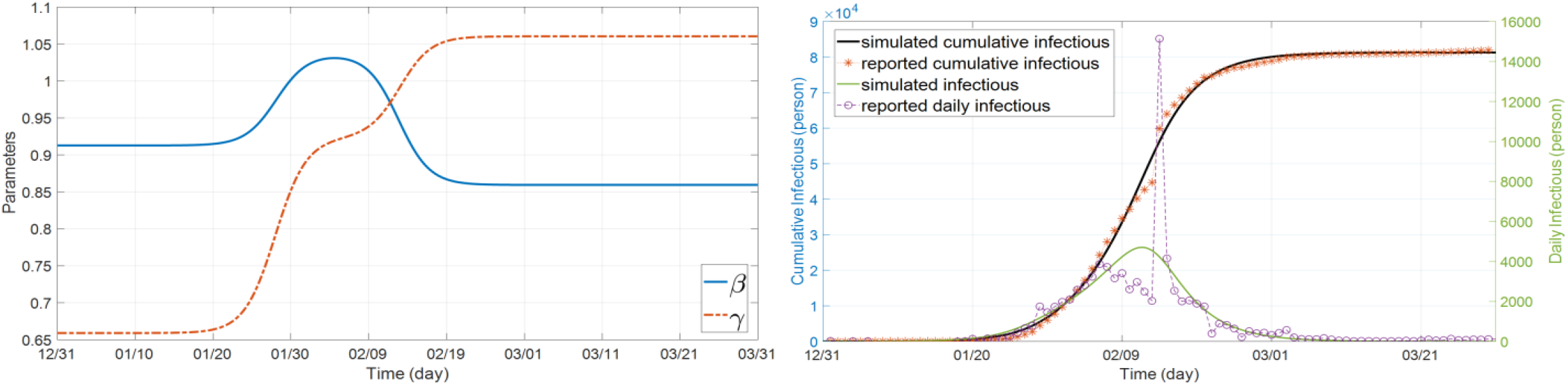
Estimated parameter values over the time using three sigmoid branches (left) and the results of infection and its cumulative plots–China outbreak

**Fig 6a.**
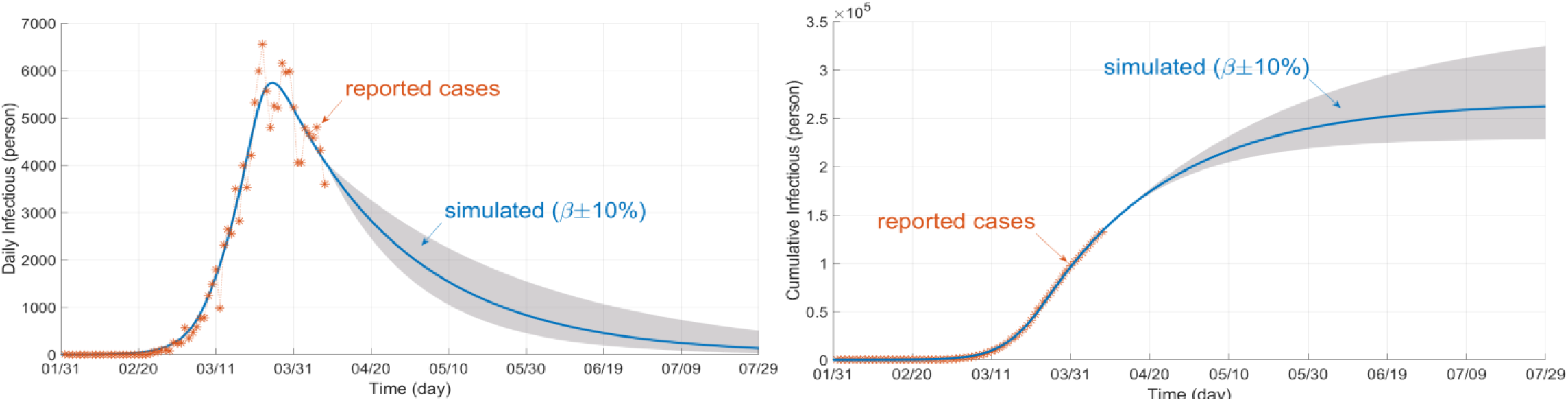
Infection plot estimated on April 7^th^, 2020 (left) and its cumulative plot (right)– 180 days period and three sigmoid branches

**Fig 6b.**
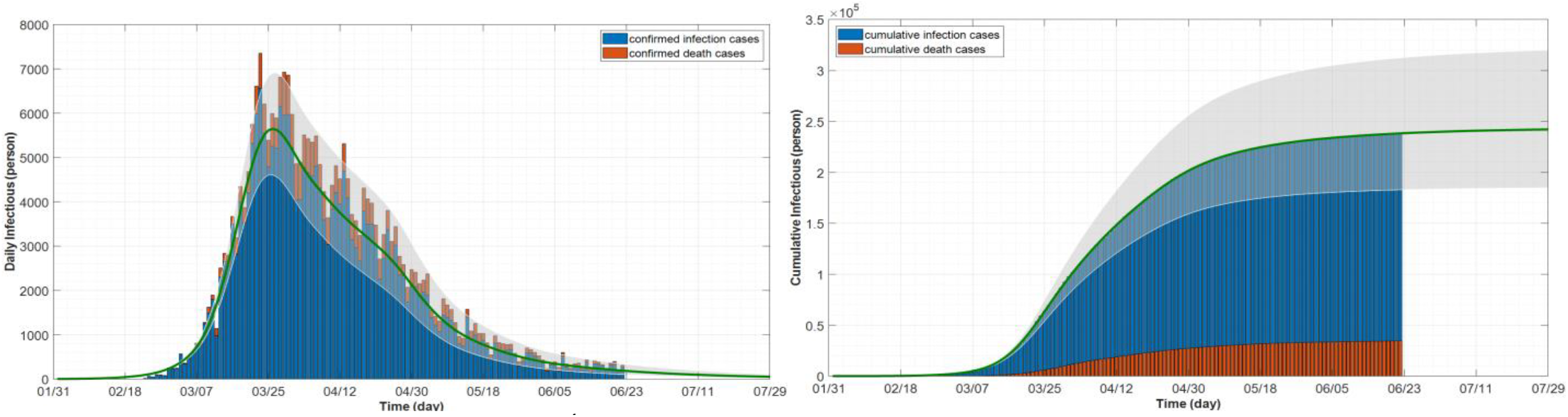
Infection plot simulated on June 22^th^, 2020 (left) and its cumulative plot (right)– 180 days period and ten sigmoid branches

**Fig 7.**
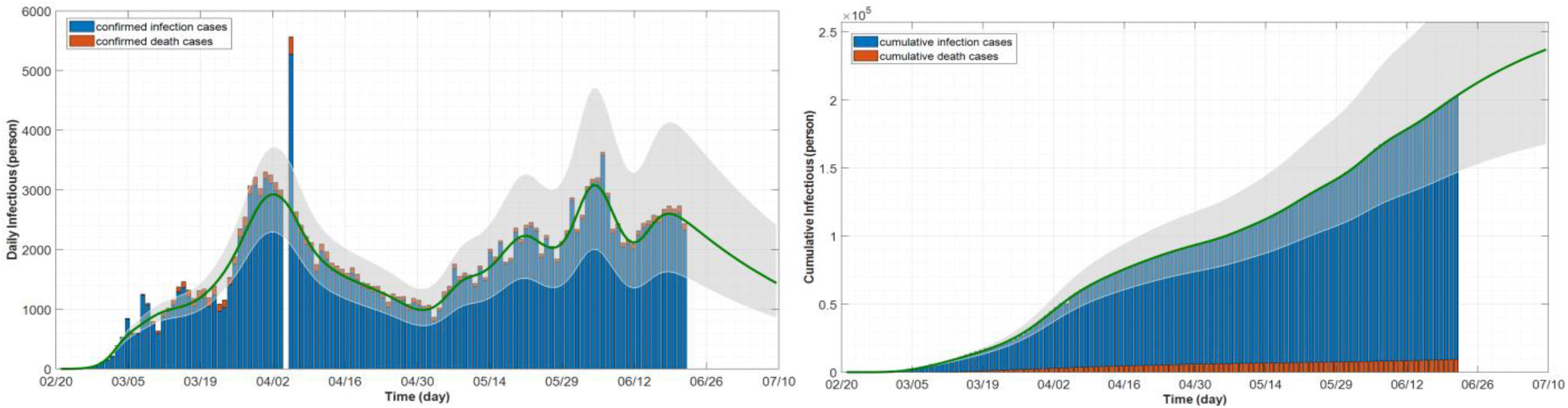
Simulation of an outbreak with multiple-dynamic changes in the reported data (including second wave of outbreak)– example of Iran, with 30 sigmoid branches

## 4. Detailed Model of SIR (SEIR, SIRD, and SEIRD)

The SIR model described above does not give information on the exposed people who are infected but not detected yet (not confirmed yet). It does not provide any knowledge of the closed cases of infectious people who have passed away. An exposed variable can be added to the SIR model to form a SEIR model, whereas a closed (death) variable can be added to the SIR model to form a SIRD model. These two variables can also be added together to the SIR model and form a new model called SEIRD. This model is more general and is adopted in this section [11]. The differential equations of this model are as follows:

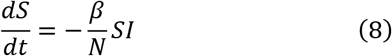

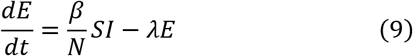

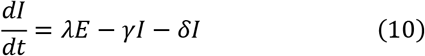

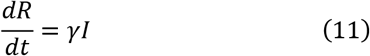

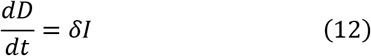

where *E* refers to the exposed state variable, *D* refers to the closed or death case, *λ* refers to the exposed parameter, and δ refers to the dead variable. Summing (8)–(12) should give zero whereas the total sum of the state variables should be constant and equals the population:

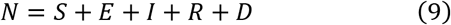

Several programs are added to SimCOVID including SEIR, SEIRD, and SEIRD designed and coded in Simulink and MATLAB. An example in Simulink is shown in Fig. 8 using the SEIRD model. Note that the actual collected data was used as input to this model. This data is stored in Excel sheets and there is a block in Simulink that allows us to import data from external sources and build a customized signal. This block is called “signal builder”. The ratio between the daily dead and daily infectious is used to represent the mortality rate. This signal represents an actual data-based signal which is multiplied by the simulated infectious signal to be integrated and form the cumulative dead variable using an integrator block in Simulink. In the figure, the orange color is used for “Exposed” and dark red is used for “Dead” compartments. The parameter optimization process and the numerical outputs are not plotted here owing to its similarity to the previous model and for space limitation reasons. However, all these models and programs are provided with this paper. The objective here in this section is to show how to edit the SIR and build a new model. Note that in the initial design of this model, a set of unit steps is used instead of sigmoid functions which can be replaced to give smoother plots.

**Fig 8.**
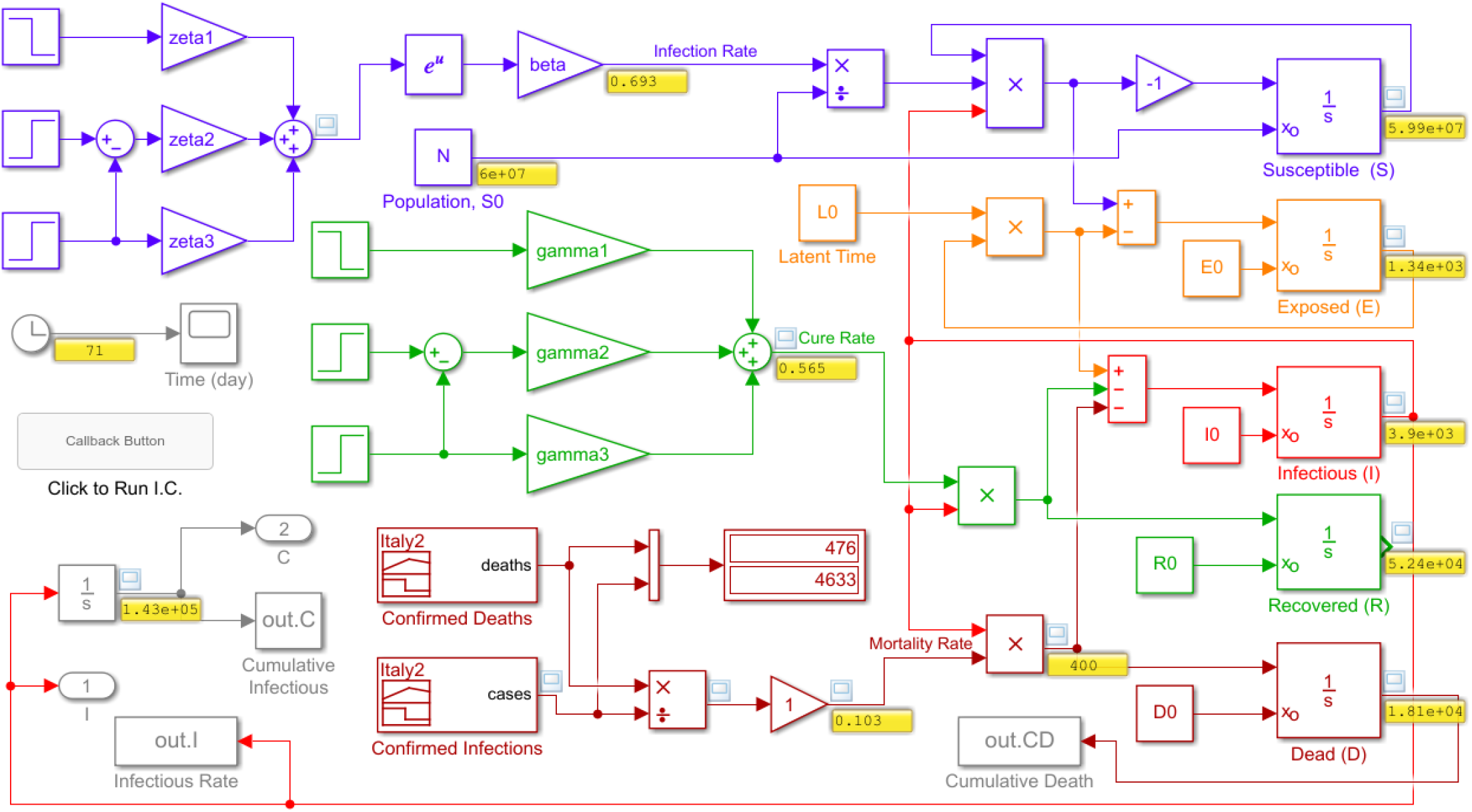
SEIRD detailed model

## 5. Simulation with control measures

Total or partial lockdown, social distancing, and stay-at-home control steps can influence the spread of the virus and flatten the curve earlier if these steps were applied in advance. In the Simulink model, we can represent all these control measures by a step function (or sigmoid function) and simulate the system under these conditions. These controls affect the infection rate giving a decline in the beta function. On the other hand, developing a vaccine or involving any other ways to recover or slow down the disease (such as providing the hospitals with all the necessary ventilators) affect the recovery-rate parameter by increasing this value. As a result, the reproduction ratio reduces by any change in these two parameters. It is also easy in Simulink to see the impact of a response delay on the curve. This can be achieved by adding a delay block to the infection function. However, more details about the model are needed for this problem.

## 6. SimCOVID Capabilities and Updates

SimCOVID went through several updates and the new version of the package is Version 5. The package started with Simulink as the platform and the first preprint was published based on this update. It was realized that some researchers like to work on MATLAB coding instead of Simulink diagrams for some reasons. For instance, unlike Simulink files, MATLAB codes can be run on different releases of the software. Another reason is that the codes can be used to build a graphical user interface. Further, the for-loop programming–which is used for the rate functions– is easier and faster in MATLAB. To keep the package useful for both researcher groups, a new set of MATLAB-based programs is added to SimCOVID. These programs have been updated several times. In the beginning, SimCOVID used three-step functions for the rate functions with a manual time setting. Step function leads to a sharp change in the simulation plots which can be avoided by replacing it with a sigmoid function providing smooth outputs. The number of sigmoid branches is also updated to a number specified by the user. Another challenge was to find the initial values of the parameters to be estimated. An update was added to automatically determine optimal initial values of the parameters that led to solve the problem even for data with multiple dynamic changes and reduce the overall simulation time. It is only required to enter the country name and run the model to simulate the outbreak for that specific country. Several statistical tools were added to choose the optimal time location for the sigmoid branches.

## 7. The educational value of SimCOVID

SimCOVID is an open-source package used for simulating, tracking, and estimating an outbreak that comes with editable files and codes. The MATLAB programs were coded in a simple way; there is only one main script for everything (reading data, parameter estimation, solving DEs, and plotting). The data itself comes from the source as an Excel sheet. A generalized method is adopted to reduce the user’s actions to solve the problem. Changing the model from one to another is also straightforward; the new equations, initial values, and their limits are appended to the existing ones.

## 8. Adaptive Neuro-Fuzzy SIR Model

The models used in the previous sections was based on the mathematical model of the problem. We can also build a machine-learning program to simulate the same system using the input and output data of the model. Simulink provides the user with an adaptive neuro-fuzzy inference system (ANFIS) toolbox to generate if-then fuzzy rules automatically based on training the given data. References [17] present a detailed description of this technique in which the same methodology is employed in this paper. Using a basic SIR model built in Simulink with a variable infection rate and constant recovery rate, the model is trained using input-out data. The input could be the infection rate, recovery rate, or a combination of the two. The output could be the infectious output or its cumulative function. In this study, the beta function and its derivative are used as input variables to the ANFIS model whereas the infectious and cumulative infectious variables are chosen for the output in two different training processes. ANFIS allows us to use only one output for each block and for this reason, two separate processes are used to generate two ANFIS blocks for the two outputs. More outputs require more ANFIS blocks.

Figure 9 shows a simple Simulink program used in this training. The recovery function is treated as constant as proposed in [6] whereas the infection rate is treated as variable [6]. With the parameters optimized, the ANFIS toolbox is used to generate seven fuzzy rules for each output. The membership function used in this training is the gaussian function. Figure 10a-c shows the training iterations, fuzzy rules, and the ANFIS outputs for the cumulative and infectious variables. Figure 11 shows the Simulink model for the ANFIS blocks used in this simulation. These if-then rules are used to simulate the case of China outbreak and the results are shown in Fig. 11a. whereas Fig. 11b shows the infections and recovery parameter values. Notably, the results show some good matching but it needs improvement. A more detailed and complicated beta function formula can be used to improve the accuracy of the results.

**Fig 9.**
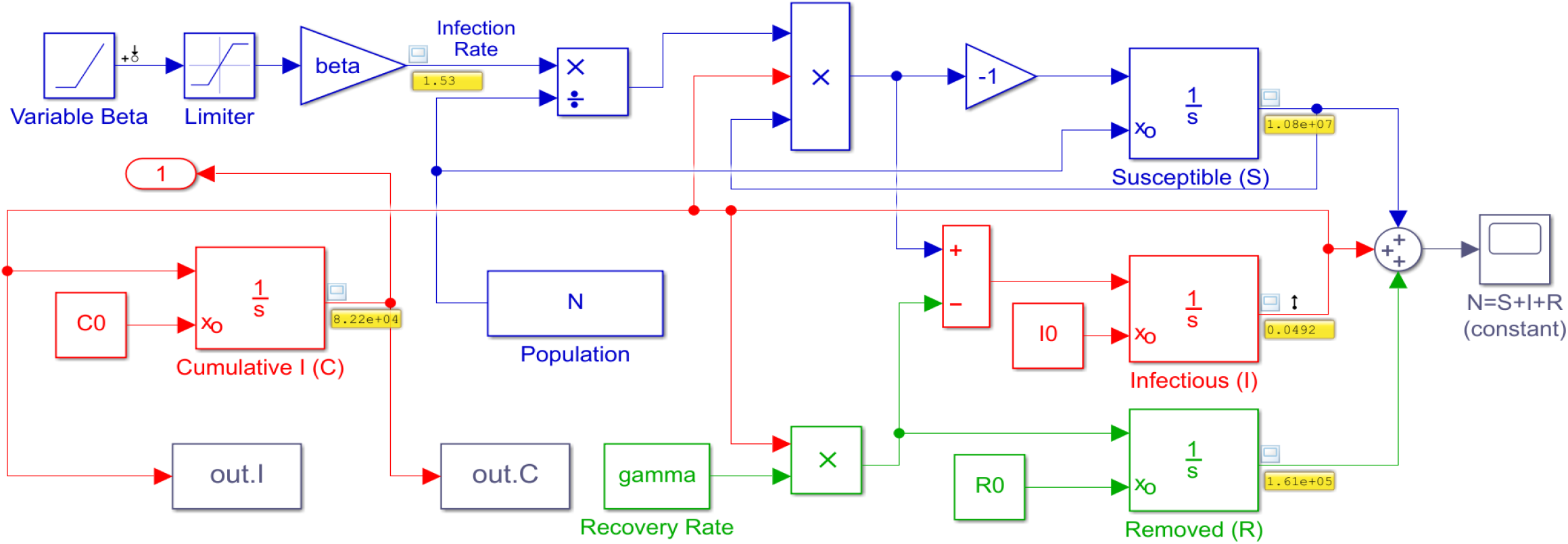
A simple Simulink program used for building an ANFIS block

**Fig 10a.**
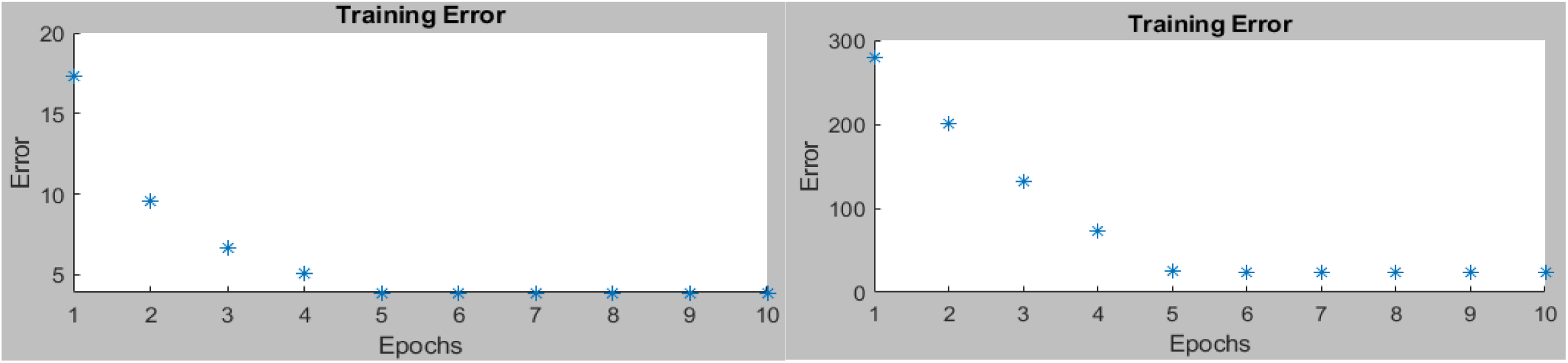
a Training errors for the infectious variable (left) and the cumulative infectious variable (right)

**Fig 10b.**
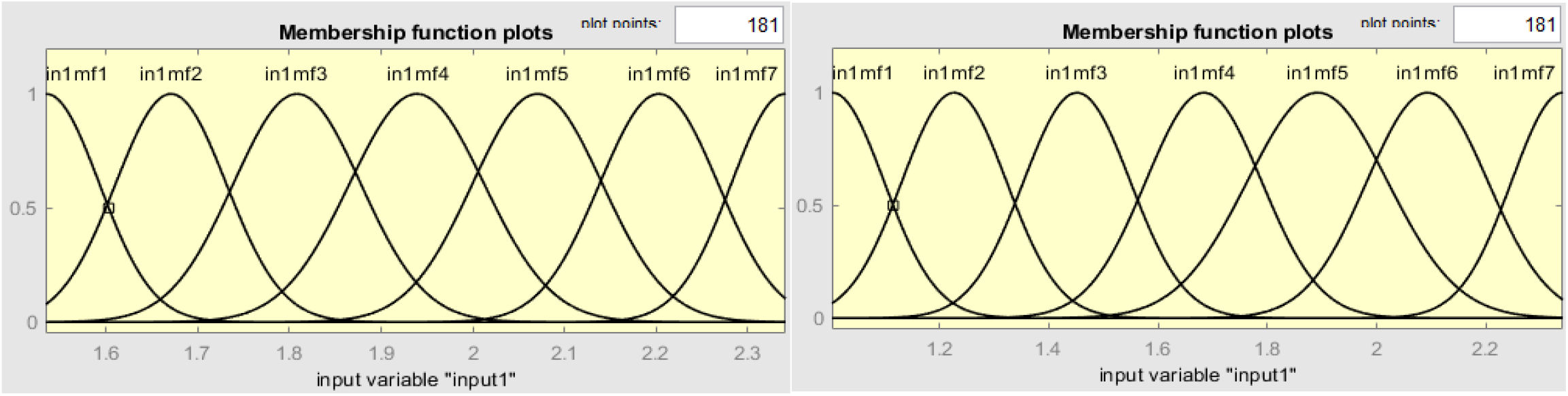
Fuzzy rules generated by ANFIS for the infectious variable (left) and the cumulative infectious variable (right)

**Fig 10c.**
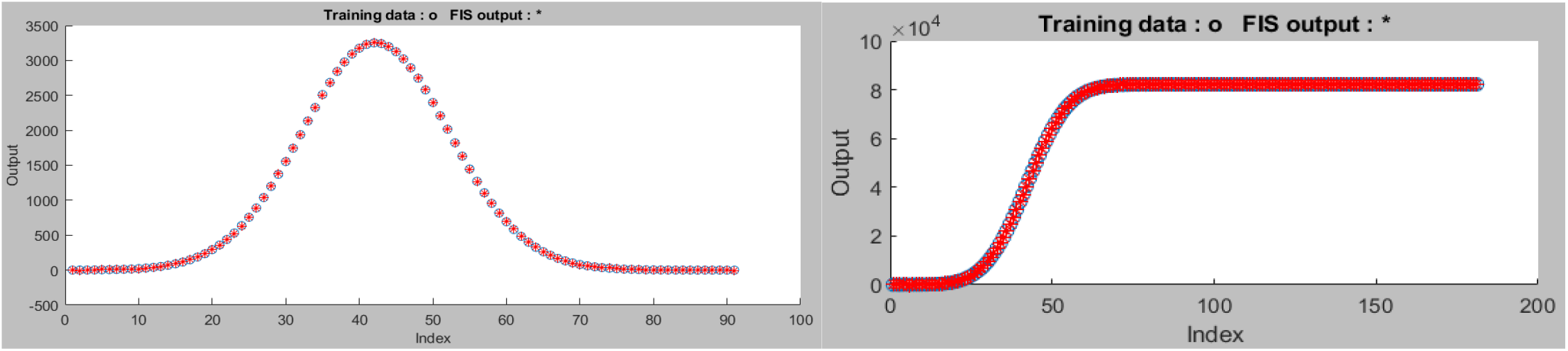
Testing data against ANFIS output for the infectious variable (left) and the cumulative infectious variable (right)

**Fig 11.**
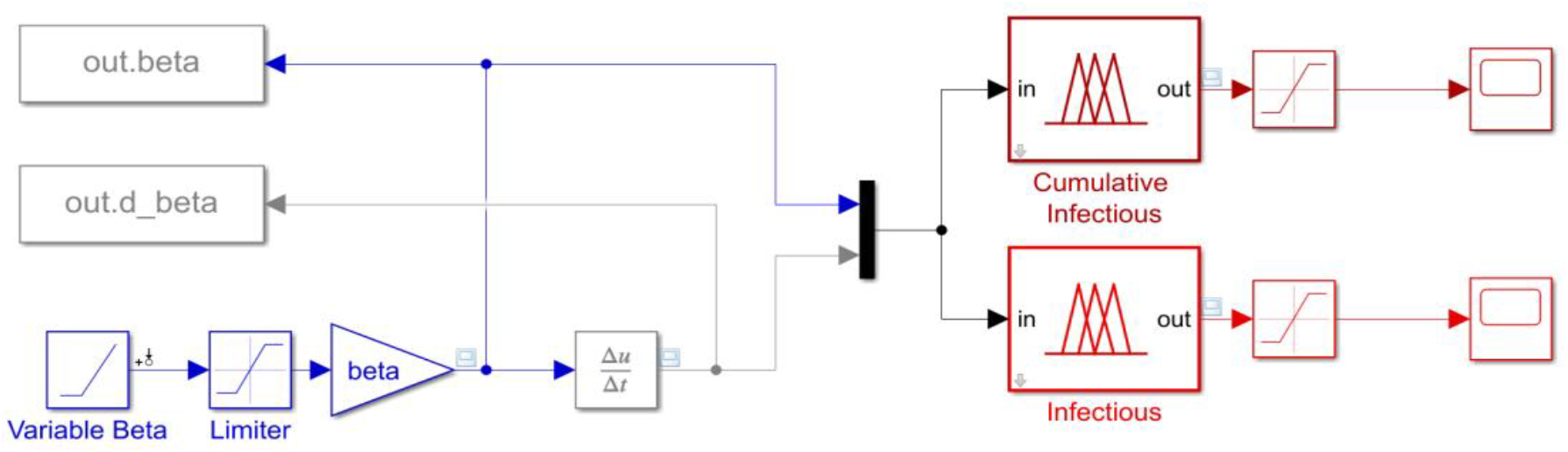
The developed ANFIS model in Simulink

**Fig 11a.**
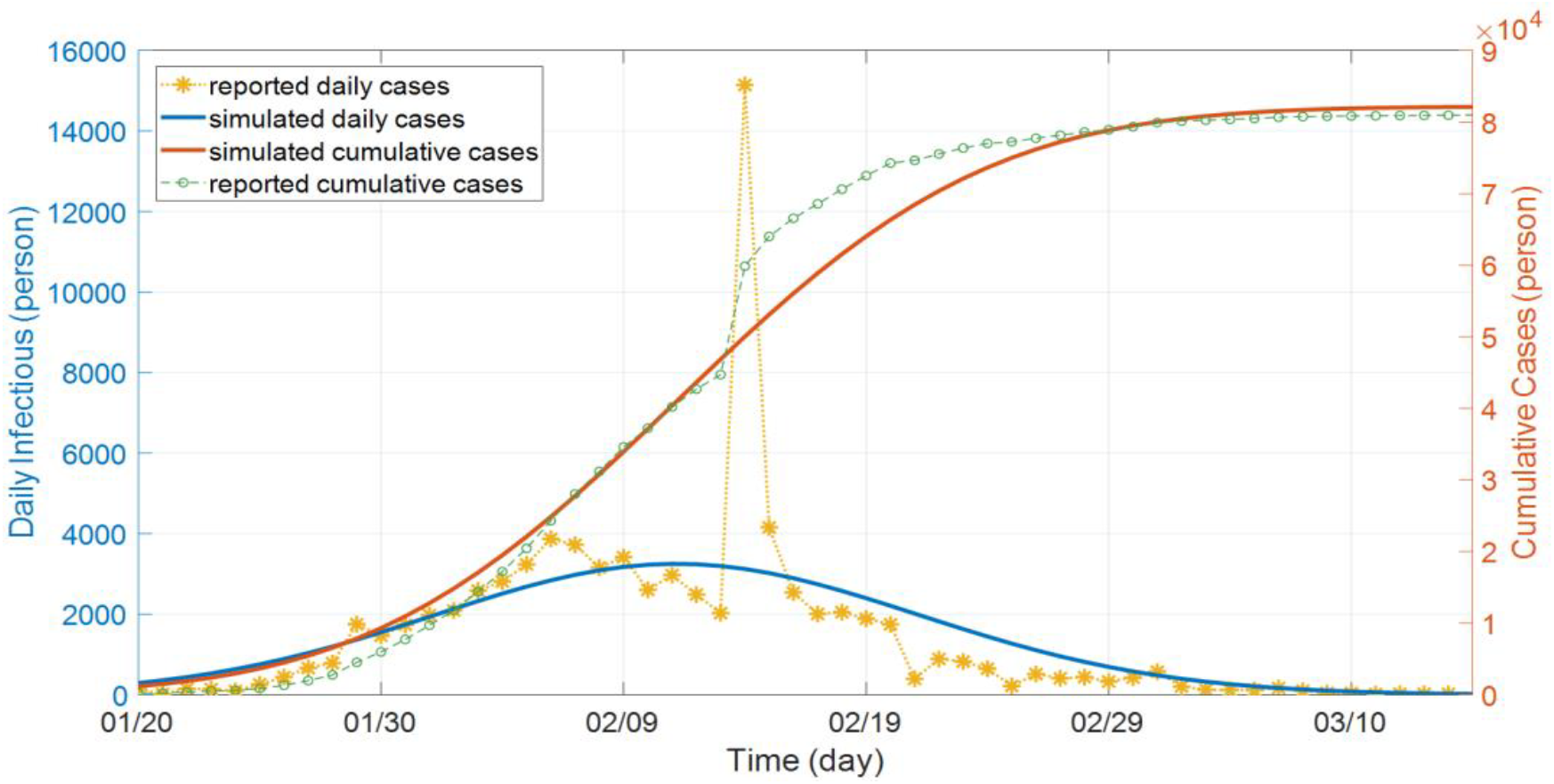
ANFIS outputs and the reported cases (daily on the left y-axis and cumulative on the right y-axis) – case of China

**Fig 11b.**
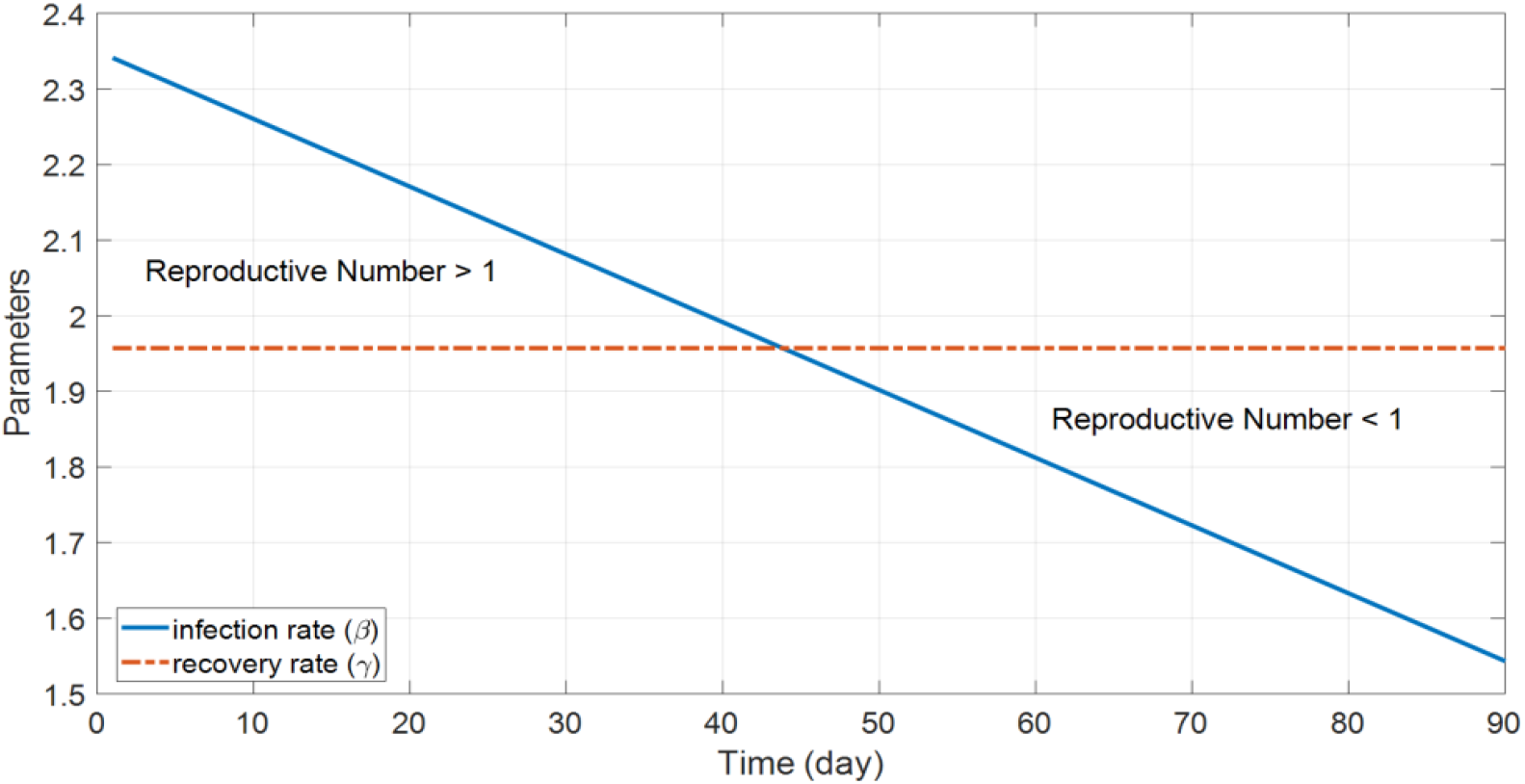
Optimized parameters used in building the ANFIS model

## 9. Conclusion

This paper presented an open-source toolbox to model, simulate, and predict the coronavirus COVID-19 outbreak using Simulink and MATLAB. The programs are easy to edit for new models, simple in their structure, generalized, fast, and can be used for simulating worldwide outbreaks with only inputting the country name. Several models were presented including SIR, SEIR, SIRD, and SEIRD models. The rate functions were treated as variables with multiple sigmoid branches to give smooth and good tracking and estimation. The initial values for the parameters to be estimated are automatically calculated based on some efficient criteria extracted from the given data. Several statistical measures were used for determining the optimal time parameters for the sigmoid functions. In addition, an adaptive neuro-fuzzy inference system was used to generate the output of the model based on some training tasks applied to the system. This paper promises some lasting values in the field of the coronavirus spread. The program can be used as an educational tool or for research studies.

## Data Availability

All data included in the paper have been referred to inside the paper (appendix).

https://www.mathworks.com/matlabcentral/fileexchange/75025-simcovid-an-open-source-simulation-program-for-the-covid-19

## Appendix I

The programs are available at: https://www.mathworks.com/matlabcentral/fileexchange/75025-simcovid-an-open-source-simulation-program-for-the-covid-19

## Appendix II

Demonstrating Videos

[1]. https://www.dropbox.com/s/sbnn1784a00hiw7/ANFIS.webm?dl=0

[2]. https://www.dropbox.com/s/mxbz8j9ogoom65g/China_Wuhan_SEIRD.webm?dl=0

[3]. https://www.dropbox.com/s/myi95d6dwnttoc8/ChinaOneStep.webm?dl=0

[4]. https://www.dropbox.com/s/mj6bi1jbiik9c03/Italy_SEIRD.webm?dl=0

[5]. https://www.dropbox.com/s/ncihp1dbqx1p35m/Italy_Optimization.webm?dl=0

[6]. https://www.dropbox.com/s/illjl95v4r603og/Italy.webm?dl=0

## References

[1]. N. Richard, D. Robert, D. Valentin, H. Emma, and A. Jan, “Potential impact of seasonal forcing on a SARS-CoV-2 pandemic”, Swiss Medical Weekly, March 2020,150:w20224. Available at: https://smw.ch/article/doi/smw.2020.20224

[2]. WHO Emergency Committee. Statement on the second meeting of the international health regulations (2005) emergency committee regarding the outbreak of novel coronavirus (2019-ncov). 2020. Available at: https://www.who.int/news-room/detail/30-01-2020-statement-on-the-second-meeting-of-the-international-health-regulations-(2005)-emergency-committee-regarding-the-outbreak-of-novel-coronavirus-(2019-ncov)

[3]. WHO Director-General’s opening remarks at the media briefing on COVID-19. 2020. Available at: https://www.who.int/dg/speeches/detail/who-director-general-s-opening-remarks-at-the-media-briefing-on-covid-19--3-april-2020.

[4]. WHO Emergency Committee. Novel Coronavirus (2019-nCoV) Situation Report – 76. 2020. Available at: https://www.who.int/docs/default-source/coronaviruse/situation-reports/20200405-sitrep-76-covid-19.pdf?sfvrsn=6ecf0977_2

[5]. Global Health NOW, Johns HopkinsBloomberg School of Publock Health, Available at: https://www.globalhealthnow.org/2020-04/1-million

[6]. L. Zhong, L. Mu1, J. Li, J. Wang, Z. Yin, and D. Liu, “Early Prediction of the 2019 Novel Coronavirus Outbreak in the Mainland China Based on Simple Mathematical Model”, DOI: 10.1109/ACCESS.2020.2979599, pp. 51761–51769, Vol. 8, 2020. Available at: https://ieeexplore.ieee.org/document/9028194

[7]. I. Nesteruk, “Statistics Based Models for The Dynamics of Chernivtsi Children Disease”, doi: DOI: 10.20535/1810-0546.2017.5.108577. Available at: http://bulletin.kpi.ua/article/view/108577

[8]. D. Caccavo1, “Chinese and Italian COVID-19 outbreaks can be correctly described by a modified SIRD model” Preprint manuscript, March 19, 2020. doi: 10.1101/2020.03.19.20039388. Available at: https://www.medrxiv.org/content/10.1101/2020.03.19.20039388v2

[9]. C. Anastassopoulou, L. Russo, A. Tsakris, C. Siettos, “Data-Based Analysis, Modelling and Forecasting of the Novel Coronavirus (2019-Ncov) Outbreak”, medRxiv preprint doi: 10.1101/2020.02.11.20022186. Available at: https://www.medrxiv.org/content/10.1101/2020.02.11.20022186v5

[10]. M. Khan, A. Atangana, Modeling the dynamics of novel coronavirus (2019-nCov) with fractional derivative, Alexandria Engineering Journal, DOI:10.1016/j.aej.2020.02.033. Available at: https://www.sciencedirect.com/science/article/pii/S1110016820300971

[11]. L. Peng et al, “Epidemic analysis of COVID-19 in China by dynamical modeling”, Preprint 2002.06563v1. Available at: https://arxiv.org/abs/2002.06563v1?utm_source=feedburner&utm_medium=feed&utm_campaign=Feed%3A+CoronavirusArXiv+%28Coronavirus+Research+at+ArXiv%29

[12]. M. Batista, Estimation of the final size of the coronavirus epidemic by the SIR model, Updated on April 06, 2020. Available at: https://www.mathworks.com/matlabcentral/fileexchange/74658-fitviruscovid19

[13]. M. Batista, Estimation of the final size of the coronavirus epidemic by the logistic model, Updated on March 25, 2020. Available at: https://www.mathworks.com/matlabcentral/fileexchange/74411-fitvirus

[14]. COVID-19 Hospital Impact Model for Epidemics (CHIME), The Trustees of the University of Pennsylvania, 2020. Available at: https://penn-chime.phl.io/

[15]. European Centre for Disease Prevention and Control, Situation update worldwide, as of 9 April 2020, Available at: https://www.ecdc.europa.eu/en/geographical-distribution-2019-ncov-cases

[16]. J. Ciarochi, How COVID-19 and Other Infectious Diseases Spread: Mathematical Modeling, March 12, 2020. Available at: https://triplebyte.com/blog/modeling-infectious-diseases

[17]. I. Abdulrahman and G. Radman, “Wide-Area-Based Adaptive Neuro-Fuzzy SVC Controller for Damping Interarea Oscillations,” in Canadian Journal of Electrical and Computer Engineering, vol. 41, no. 3, pp. 133–144, Summer 2018. Available at: https://ieeexplore.ieee.org/document/8506642

